# Plasma brain-derived p-Tau217 outperforms other p-Tau species in detecting abnormal brain amyloid in an Asian cohort of older people with cerebrovascular disease burden

**DOI:** 10.64898/2025.12.09.25341888

**Authors:** Joyce R. Chong, Saima Hilal, Narayanaswamy Venketasubramanian, Michael Schöll, Nicholas J. Ashton, Henrik Zetterberg, Christopher P. Chen, Mitchell K. P. Lai

## Abstract

**INTRODUCTION:** We evaluated the performance of plasma brain-derived (BD)- as well as total-p-Tau181, p-Tau217 and p-Tau231 in detecting beta-amyloid positivity (Aβ+) and cognitive decline in a Singapore-based cohort of older people with cerebrovascular disease.

**METHODS:** Brain amyloid status (Aβ- [n = 139] vs Aβ+ [n = 74]) was determined by positron emission tomography (PET) scans. Plasma BD and total p-Tau were measured using NUcleic acid Linked Immuno-Sandwich Assay multiplexing platform (NULISAseq).

**RESULTS:** BD-p-Tau217 (area under the curve [AUC] = 0.965) outperformed other BD and total-p-Tau species in detecting PET Aβ+ (AUC = 0.823-0.937; all p ≤ 0.008). Using three-range or binary references, BD-p-Tau217 demonstrated high sensitivity (>90%), specificity (>90%), positive (>85%) and negative (>95%) predictive values. BD-p-Tau217-derived High-risk group exhibited faster cognitive decline than the Low-risk group.

**DISCUSSION:** Risk stratification for PET Aβ+ based on plasma BD-p-Tau217 suggests superior diagnostic and prognostic utility, warranting further assessment.

## 1. INTRODUCTION

Blood-based biomarkers of brain amyloid burden, particularly those derived from phosphorylated Tau (p-Tau), are gaining recognition for their clinical utility and potential for use in routine assessments for Alzheimer’s disease (AD) [1]. Clinically significant p-Tau species, including p-Tau181, p-Tau217 and p-Tau231 are now quantifiable via a range of assays and platforms [1, 2], and blood p-Tau biomarkers correlate well with positron emission tomography (PET) measures of brain amyloid or AD cerebrospinal fluid biomarkers [2–5]. From a comprehensive systematic review evaluating the diagnostic performance of plasma p-Tau biomarkers and associated assays [1], as well as a recent study comparing 33 different plasma p-Tau assays [2], plasma p-Tau217 has emerged as the best-performing biomarker for identifying AD pathology, showing the highest areas under the receiver-operating characteristic curve (AUC) and the largest fold changes compared with other p-Tau species, with broadly similar performance across different p-Tau217 assays. Notably, with the growing availability of commercial plasma p-Tau217 assays, a slew of recent studies have demonstrated the diagnostic utility of this biomarker for detecting abnormal brain amyloid burden in both Western [6–11] and Asian [12–16] populations, supporting its accessibility and generalizability across different populations. However, one important caveat with currently available, widely used p-Tau assays is that most of them utilize detection antibodies that recognize an epitope common to both brain-derived (BD), low-molecular-weight (LMW) Tau, as well as high-molecular-weight (HMW) Tau produced by the peripheral nervous system (PNS) and other organs, in effect measuring “total-Tau” [2, 17]. Briefly, HMW Tau (110 kDa) contains additional exons, including the 250-amino acid exon 4a [18]. In contrast, all six LMW isoforms (37 kDa to 46 kDa) in the brain exclude exon 4a [18, 19]. As an illustration of the potential significance of differentiating BD from peripheral Tau isoforms, recent studies measuring total p-Tau have reported increased blood p-Tau217 and p-Tau181 in patients with amyotrophic lateral sclerosis (ALS). Assessment of p-Tau immunoreactivities in muscle biopsies suggested that the increased p-Tau may reflect changes in atrophic muscle fibers in ALS, and was independent of AD pathology in the brain [20, 21]. In this context, assays which specifically measure plasma LMW BD-p-Tau species may be able to mitigate the confounding effect of peripheral p-Tau, thereby improving sensitivity and specificity.

In this study, using a Singapore-based cohort of longitudinally assessed older people with high baseline cerebrovascular disease burden, we employed a NUcleic acid Linked Immuno-Sandwich assay (NULISAseq) with antibodies that preferentially bind LMW Tau, or bind the common region of both LMW and HMW Tau, coupled with those specific for each phospho-epitope, in order to simultaneously measure BD-p-Tau217, BD-p-Tau181, and BD-p-Tau231, as well as their corresponding total-p-Tau species. We then performed head-to-head comparisons of plasma BD-p-Tau217 with the other p-Tau species for the detection of brain amyloid burden (Aβ+) on positron emission tomography (PET). Furthermore, we evaluated the references ranges for abnormal amyloid burden and assessed the prognostic performance of the plasma BD-p-Tau217 reference ranges.

## 2. MATERIALS AND METHODS

### 2.1 Study population

HARMONISATION, a Singapore-based prospective study, enrolled 700 participants from the National University Hospital and St Luke’s Hospital Memory Clinics, as well as the community, with annual follow-up for up to 5 years [22]. Of the participants, 217 individuals were also recruited for amyloid positron emission tomography (PET) assessment [12, 23–25]. Participants were classified into one of the following diagnostic categories: (1) cognitively normal (CN): individuals without objective cognitive impairment on formal neuropsychological assessments, or functional loss; (2) cognitive impairment no dementia (CIND): individuals with impairment in at least one cognitive domain on neuropsychological assessments, but who did not meet the Diagnostic and Statistical Manual of Mental Disorders, Fourth Edition (DSM-IV) criteria for dementia; (3) dementia: individuals diagnosed according to the DSM-IV criteria, with further etiologic diagnoses following the National Institute of Neurological and Communicative Disorders and Stroke and the Alzheimer’s Disease and Related Disorders Association (NINCDS-ADRDA) criteria for AD, and the National Institute of Neurological Disorders and Stroke and Association Internationale pour la Recherché et l’Enseignement en Neurosciences (NINDS-AIREN) criteria for vascular dementia (VaD). At baseline, participants underwent medical history taking and clinical interviews, including the assessment of vascular risk factors (hypertension, hyperlipidaemia, diabetes mellitus, and cardiovascular disease). They also underwent blood collection, Apolipoprotein E (*APOE*) genotyping, and brain magnetic resonance imaging (MRI) to assess significant cerebrovascular disease (CeVD+), defined as the presence of a total number of cortical infarcts ≥1, and/or total number of lacunes ≥2, and/or total number of cerebral microbleeds ≥2, and/or Fazekas score ≥2 [26]. Cognitive assessments, including Mini-Mental State Examination (MMSE), Clinical Dementia Rating Sum-of-Boxes (CDR-SB), and a comprehensive neuropsychological battery consisting of seven cognitive domains (Executive Function, Attention, Language, Visuomotor Speed, Visuoconstruction, Verbal Memory, and Visual Memory), were administered as previously described [12, 23–25]. Clinical diagnosis, blood collection, MMSE, and CDR-SB assessments were repeated annually for up to 5 years.

Of the 217 participants recruited for the amyloid PET study, 213 also had plasma NULISAseq measurements, with a mean (SD) interval of 35 (24) months between blood collection and PET imaging. **Table 1** presents demographics and clinical characteristics for these 213 participants, using data from the blood collection visit (age, sex, years of education, *APOE4* carrier status, MMSE, clinical diagnosis) and at start of recruitment (vascular risk factors and CeVD status).

**Table 1.**
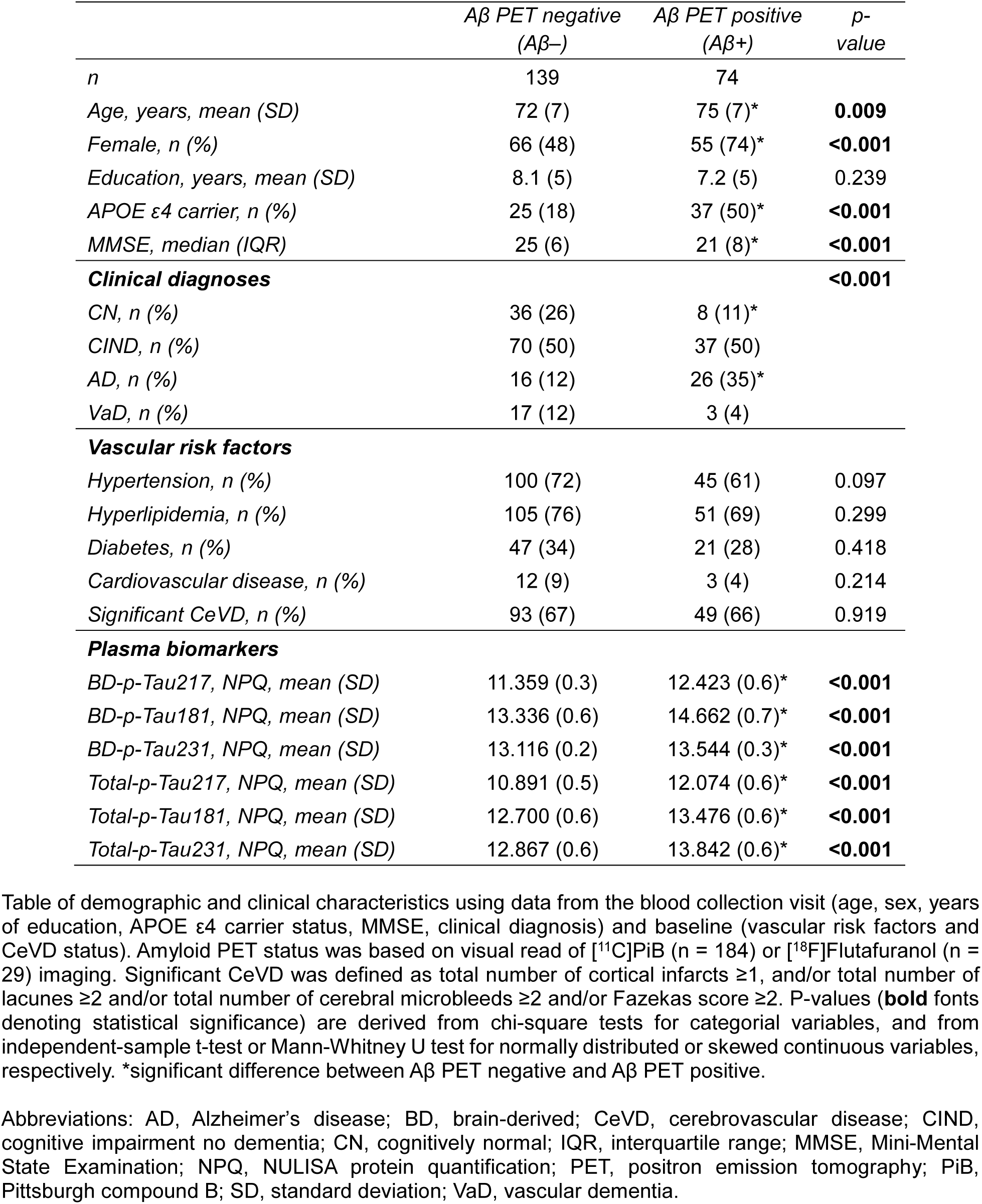
Characteristics of study participants.

### 2.2 NULISAseq analysis and data processing

Non-fasting blood was collected into ethylenediaminetetraacetic acid (EDTA)–containing tubes and centrifuged at 2000 *g* for 10 minutes at 4◦C. Plasma was extracted, mixed well, aliquoted and stored at −80◦C until use. NULISAseq assays were performed with evaluators blinded to clinical information at Alamar Biosciences Inc (Fremont, CA), as described previously [27]. Briefly, plasma samples were thawed on ice and centrifuged at 2200 *g* for 10mins, after which 25uL supernatant aliquots were run on the ARGO™ HT multiplexing platform using the NULISAseq™ CNS Disease Panel 120, starting with immunocomplex formation with pairs of DNA barcode-containing, poly adenine nucleotide- or biotin-conjugated capture and detection antibodies, followed by sequential capturing and washing on paramagnetic oligo-dT beads, then on streptavidin beads.

Finally, the proximal ends of the DNA strands on the retained detection and capture antibody complexes were ligated to generate a DNA reporter molecule containing both target-specific and sample-specific barcodes. DNA reporter molecules were pooled and amplified by polymerase chain reaction (PCR), purified and sequenced on Illumina NextSeq 2000 next generation sequencer. Sequencing data were processed using the NULISAseq algorithm (Alamar Biosciences) to quantify the sample- (SMI) and target-specific (TMI) barcodes (molecular identifiers), and intraplate normalization was performed by dividing the target counts for each sample well by that well’s internal control counts. Interplate normalization was then performed using interplate control (IPC) normalization, wherein counts were divided by target-specific medians of the three IPC wells on that plate. Data were then rescaled, added 1, and log2 transformed to obtain NULISA Protein Quantification (NPQ) units for downstream statistical analysis (see **Figure 1A** for a summary of workflow). Specificity for BD (LMW) Tau isoforms was achieved with antibodies which recognize an epitope at the junction between MAPT exons 4 and 5 (thus excluding HMW isoforms which have exons 4 and 4a junctions), while antibodies which recognize the common N terminal epitope will detect both LMW and HMW isoforms. These antibodies are paired with antibodies specific for phospho-epitopes at threonine-181, 217 or 231, thus enabling the measurements of BD- and total-p-Tau species (see **Figure 1B**).

**Figure 1.**
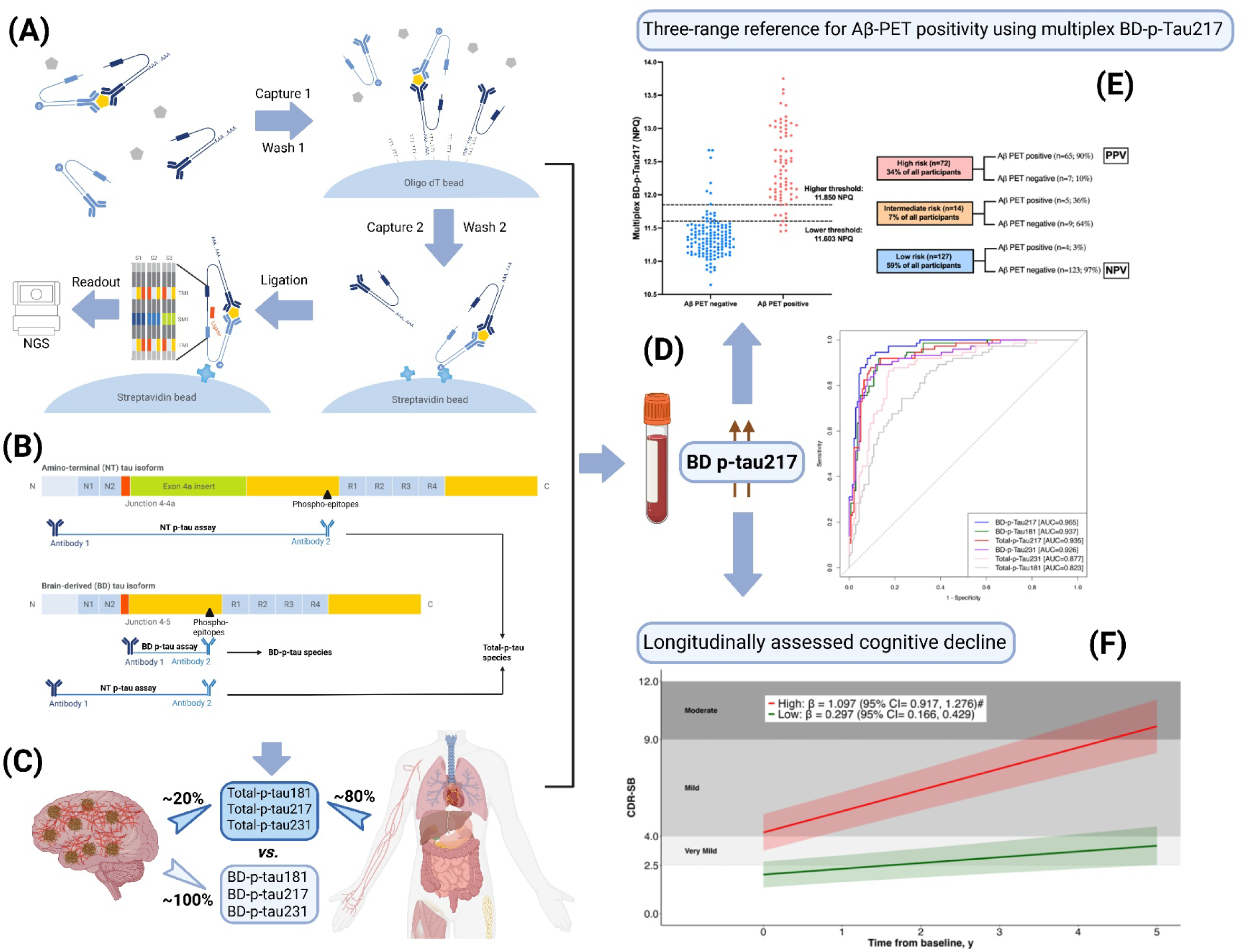
Graphical summary of study workflow and major findings. (A) Principle of NULISAseq, consisting of DNA-barcode-containing detection-capture antibody pairs, one conjugated with poly-adenine nucleotides and the other with biotin. Each pair of antibodies recognize different epitopes on the same target molecule, forming a tripartite immune complex, which are then sequentially captured and washed on oligo-dT beads (“Capture 1 / Wash 1”), then on streptavidin beads (“Capture 2 / Wash 2”), with the proximal ends of the DNA strands of the retained, complexed antibodies ligated to generate a DNA reporter molecule containing both target-specific and sample-specific barcodes. DNA reporter molecules were pooled and amplified by polymerase chain reaction, purified and sequenced on a Next-Generation sequencer (NGS), with sequencing data processed by NULISAseq algorithm for the quantification of sample- (SMI) and target-specific (TMI) barcodes (molecular identifiers) to derive NULISA Protein Quantification (NPQ) units for downstream statistical analysis. (B) Specificity for BD (LMW) Tau isoforms was achieved with antibodies which recognize an epitope at the junction between MAPT exons 4 and 5, while antibodies which recognize the common amino terminus (NT) region will detect both LMW and HMW isoforms. These antibodies are paired with antibodies specific for phospho-epitopes at threonine-181, 217 or 231, thus enabling the measurements of BD- and total-p-Tau species. (C) Postulated sources of BD- and total-p-Tau species. (D) AUC showing the superior utility of BD-p-Tau217 to (E) stratify amyloid risk based on a three-range reference and (F) predict for longitudinal cognitive impairment. Abbreviations: AUC, area under the receiver-operating characteristic (ROC) curve; BD, Brain-derived; CDR-SB, Clinical Dementia Rating Sum of Boxes; NGS, Next-Generation sequencer; NPV, Negative predictive value; NT, Amino terminal; NULISAseq, NUcleic acid Linked Immuno-Sandwich Assay multiplexing platform; PET, Positron emission tomography; PPV, Positive predictive value; SMI, Sample-specific molecular identifier; TMI, Target-specific molecular identifier.

### 2.3 Simoa p-Tau217 measurements

In a subset of 152 participants, plasma Simoa p-Tau217 levels were measured using the commercial ALZpath p-Tau217 Advantage PLUS assay (#104570) on the HD-X platform (Quanterix Corp., Billerica, MA) as previously described [12]. Plasma Simoa p-Tau217 were measured from blood collected during the same visit as the NULISA CNS sample. Evaluators were blinded to clinical information and the immunoassays were performed following the manufacturer’s protocol. Both inter- and intra-assay coefficients of variation were <10%.

### 2.4 Amyloid PET acquisition and quantification

Details on the amyloid PET acquisition and quantification were as previously described [12, 23–25]. Briefly, 184 participants underwent a 30-min brain PET scan after injection of 370 MBq of [^11^C]-Pittsburgh Compound B (PiB) on an mMR synchronous PET/MR scanner (Siemens Healthcare GmbH, Munich, Germany). An additional 29 participants underwent a 20-min brain PET scan on a mCT PET-CT (computed tomography) scanner (Siemens Healthcare GmbH), after intravenous injection of 185 MBq of [^18^F]-flutafuranol. Images were reconstructed using ordinary Poisson ordered-subsets expectation maximization with all corrections applied. Each [^11^C]-PiB or [^18^F]-flutafuranol image was independently visually interpreted by three experienced raters blinded to the clinical diagnosis of each participant and was used to classify individuals as Aβ positive (Aβ+) or Aβ negative (Aβ-) following established criteria. Global standardized uptake ratio (SUVr) values were derived from the [^11^C]-PiB scans and individually parcellated MRI for reference and target region definition, using an in-house developed automated pipeline.

### 2.5 Statistical analyses

Statistical analyses were performed using SPSS version 29 (IBM Corp., Armonk, NY) and R version 4.4.0 (R Project for Statistical Computing). For the comparison of characteristics between PET Aβ- and Aβ+ groups, independent-sample t-tests or Mann–Whitney U tests were used for normally distributed or skewed continuous variables, respectively; while chi-square tests were used for categorical variables. Differences in plasma biomarkers among groups stratified by clinical diagnosis (CN, CIND, dementia) and Aβ-PET status (Aβ-, Aβ+) were assessed using the Kruskal-Wallis test followed by *post-hoc* Dunn’s test with Bonferroni correction. Correlations were examined using the Spearman’s correlation, with bootstrapping (n = 2000 iterations) used to test differences in the correlation coefficients. Diagnostic performance for detecting Aβ-PET positivity was assessed using the area under the receiver-operating characteristic curve. AUC and 95% confidence-intervals (CIs) were computed using DeLong’s method with the pROC package [28]. The p-values for comparisons of AUCs were derived from DeLong tests. To determine the reference ranges for Aβ-PET positivity, we used a three-range strategy comprising a lower reference point to rule out AD (95% sensitivity; “Low-risk” for PET Aβ+) and a higher reference point to rule in AD (95% specificity; “High-risk” for PET Aβ+) [6, 29, 30]. Participants with values between the reference points were classified as having “Intermediate risk” which constituted the group requiring confirmatory Aβ PET testing. Alternatively, a binary reference point for Aβ-PET positivity was derived based on maximizing the Youden index [6]. We evaluated the concordance of a low-risk result with Aβ-PET negativity (negative predictive value [NPV]), as well as the concordance of a high-risk result with Aβ-PET positivity (positive predictive value [PPV]). To assess the associations between plasma BD-p-Tau217-derived risk groups with baseline and longitudinal cognitive performance, linear mixed-effects (LME) models were used with MMSE or CDR-SB as outcome, including as predictors the plasma BD-p-Tau217-derived risk groups, time, age, sex and years of education, as well as an interaction between risk groups and time. The model contained random intercepts and random slopes, and time was modeled as a continuous variable. Plasma BD-p-Tau217-derived risk groups were entered as dichotomous variable (Group 1: Low-risk, Group 2: High-risk). To assess the associations between plasma BD-p-Tau217-derived risk groups and risk of progression to dementia, we used Cox proportional hazards regression models, adjusting for age, sex and education. For all analyses, p-values < 0.05 were considered statistically significant.

## 3. RESULTS

### 3.1 Participants characteristics

Demographic and clinical information, as well as plasma biomarker measurements, are listed in **Table 1**. A total of 213 participants were included in this study (mean [SD] age 73 [7] years; 121 females [57%]; 44 CN [21%], 107 CIND [50%], 42 AD [20%] and 20 VaD [9%]). Compared with the PET Aβ- participants, the PET Aβ+ participants were older, had a higher proportion of females, a higher prevalence of *APOE4* carriers and clinical AD, and lower MMSE scores (all p < 0.05). There was no difference between Aβ- and Aβ+ participants in the prevalence of vascular risk factors, including hypertension, hyperlipidaemia, diabetes and cardiovascular disease, as well as CeVD burden. Notably, both groups had high baseline CeVD burden (proportion of significant CeVD: Aβ- = 67%; Aβ+ = 66%), in agreement with previous findings from this cohort [12, 23]. For plasma biomarkers, all the BD-p-Tau (BD-p-Tau217, BD-p-Tau181 and BD-p-Tau231) and total-p-Tau (total-p-Tau217, total-p-Tau181 and total-p-Tau231) species were significantly higher in the Aβ+ participants (all *p* < 0.001).

All plasma BD- and total-p-Tau species positively correlated with PiB-PET SUVr (**Supplementary Figure S1**; Spearman’s rho = 0.488 to 0.722, all *p* < 0.001), with BD-p-Tau217 showing the highest correlation (Spearman’s rho = 0.722). Interestingly, all BD-p-Tau species showed stronger correlation with PiB-PET SUVr than their respective total-p-Tau species (BD-p-Tau217 vs. total-p-Tau217: difference in Spearman’s rho = 0.04, 95% CI = -0.02 to 0.10, *p* = 0.14; BD-p-Tau181 vs. total-p-Tau181: difference in Spearman’s rho = 0.17, 95% CI = 0.09 to 0.26, *p* <0.001; BD-p-Tau231 vs. total-p-Tau231: difference in Spearman’s rho = 0.08, 95% CI = 0.007 to 0.15, *p* = 0.025).

### 3.2 Levels of plasma BD- and total-p-Tau species stratified by clinical diagnosis and brain amyloid status

When stratified by clinical diagnosis (CN, CIND, dementia) and brain amyloid status (Aβ-, Aβ+) (**Supplementary Figure S2**), all plasma BD-p-Tau species were significantly higher in the Aβ+ participants compared with the Aβ-participants, across all diagnostic groups (all *p* ≤ 0.020). Corroborating our previous findings using a different assay platform (Simoa) [12], among the total-p-Tau species, only total-p-Tau217 showed a significant difference between CN Aβ- and CN Aβ+ participants (*p* = 0.034 for total-p-Tau217, *p* = 1.00 and 0.30 for total-p-Tau181 and total-p-Tau231, respectively). In the CIND and dementia groups, all plasma total-p-Tau species were significantly higher in the Aβ+ participants as compared with the Aβ-participants.

Among the BD-p-Tau species, BD-p-Tau181 showed the largest fold-changes between Aβ- and Aβ+ in all clinical stages (CN [percentage increase for BD-p-Tau181; BD-p-Tau217; BD-p-Tau231 respectively]: 96%; 65%; 17%, CIND: 115%; 77%; 27%, and dementia: 197%; 123%; 40%). In contrast, total-p-Tau217 showed the largest fold-changes amongst the total-p-Tau species between Aβ- and Aβ+ (CN [percentage increase for total-p-Tau217, total-p-Tau231 and total-p-Tau181 respectively]: 82%; 37%; 24%, CIND: 118%; 93%; 78% and dementia: 142%; 112%; 99%).

### 3.3 Diagnostic performance of BD- and total-p-Tau species in identifying abnormal brain amyloid burden

We next evaluated the diagnostic performance of the plasma BD- and total-p-Tau species for identifying abnormal Aβ PET. **Table 2** showed that plasma BD-p-Tau217 demonstrated superior diagnostic accuracy, achieving AUC of 0.965 (95% CI = 0.942 to 0.987) and outperforming all other BD- and total-p-Tau species (BD-p-Tau181: AUC = 0.937; total-p-Tau217: AUC = 0.935; BD-p-Tau231: AUC = 0.926; total-p-Tau231: AUC = 0.877; total-p-Tau181: AUC = 0.823; all *p* ≤ 0.008). Due to the relatively wide interval between blood collection and PET, sensitivity analyses were performed in a subset of participants (n=114) with amyloid PET within three years of blood collection, which showed similar results (**Supplementary Table S1**).

**Table 2.** Diagnostic performance of the brain-derived and total p-Tau species in identifying abnormal brain amyloid burden.

| *PET Aβ+ = 74,*<br>*PET Aβ- = 139* | *AUC*<br>*(95% CI)* | *p-value when*<br>*compared to*<br>*BD-p-Tau217* |
| --- | --- | --- |
| *BD-p-Tau217* | 0.965<br>(0.942, 0.987) | NA |
| *BD-p-Tau181* | 0.937<br>(0.904, 0.969) | **0.004** |
| *Total-p-Tau217* | 0.935<br>(0.901, 0.969) | **0.003** |
| *BD-p-Tau231* | 0.926<br>(0.889, 0.964) | **0.008** |
| *Total-p-Tau231* | 0.877<br>(0.828, 0.926) | **<0.001** |
| *Total-p-Tau181* | 0.823<br>(0.766, 0.879) | **<0.001** |

AUC and the associated 95% CI for each of the plasma BD-p-Tau (BD-p-Tau217, BD-p-Tau181, BD-p-Tau231) and total-p-Tau (total-p-Tau217, total-p-Tau181, total-p-Tau231) species, in predicting Aβ-PET positivity. Plasma BD-p-Tau217 outperformed other BD-p-Tau and total-p-Tau species in detecting abnormal brain amyloid burden ( $p \leq 0.008$ ). P-values (**bold** fonts denoting statistical significance) for pairwise comparisons of AUCs were derived from DeLong tests.
Abbreviations: AUC, area under the receiver-operating characteristic (ROC) curve; BD, brain-derived; CI, confidence interval; PET, positron emission tomography

Additionally, in a subset of participants with available Simoa plasma p-Tau217 measurements, we performed head-to-head comparisons of the NULISA measurements with Simoa p-Tau217 (n=152). **Supplementary Table S2** showed that plasma BD-p-Tau217 (AUC = 0.974, 95% CI = 0.953 to 0.996) was superior to Simoa p-Tau217 (AUC = 0.917, 95% CI = 0.864 to 0.970, *p* = 0.007) in detecting PET Aβ+. Simoa p-Tau217 still outperformed NULISAseq total-p-Tau231 (AUC = 0.881, *p* = 0.024) and total-p-Tau181 (AUC = 0.833, *p* = 0.001), whilst performing similarly with NULISAseq total-p-Tau217 (AUC = 0.947, *p* = 0.094), BD-p-Tau181 (AUC = 0.943, *p* = 0.155) and BD-p-Tau231 (AUC = 0.938, *p* = 0.255).

Finally, we examined potential effects of CeVD burden on the diagnostic performance of NULISAseq BD-p-Tau217. When stratified by CeVD status, the CeVD-group showed higher AUC (0.994, 95% CI = 0.983 to 1.00; *p* = 0.02), though AUC in the CeVD+ group remained high (0.952, 95% CI = 0.918 to 0.985). With AUC values of >0.95 in CeVD- and CeVD+ subgroups, our data suggest the suitability for use of BD-p-Tau217 in Asian populations with concomitant CeVD burden.

### 3.4 Three-range reference of plasma BD- and total-p-Tau species for abnormal amyloid pathology

To determine the three-range reference for PET Aβ+, we created lower (95% sensitivity) and upper (95% specificity) reference points for each plasma p-Tau species (**Table 3**, **Figure 2A** and **Supplementary Figure S3**). For plasma BD-p-Tau217 (lower threshold: BD-p-Tau217 ≤11.603 NPQ; higher threshold: BD-p-Tau217 ≥11.850 NPQ), this approach resulted in NPV and PPV of 97% and 90% respectively. The proportion of participants in the intermediate-risk group, who would in practice be followed-up for confirmatory Aβ PET, was 7%. In contrast, while the other plasma BD- and total-p-Tau species achieved largely similar NPVs (94% to 96%) and PPVs (77% to 89%), the proportion of participants in the intermediate-risk group was substantially higher (20% to 57%). For example, plasma total-p-Tau217 (lower threshold: total-p-Tau217 ≤11.082 NPQ; higher threshold: total-p-Tau217 ≥11.729 NPQ) yielded NPV and PPV of 96% and 89% respectively. However, the proportion of participants in the intermediate-risk group was 23%.

**Table 3.**
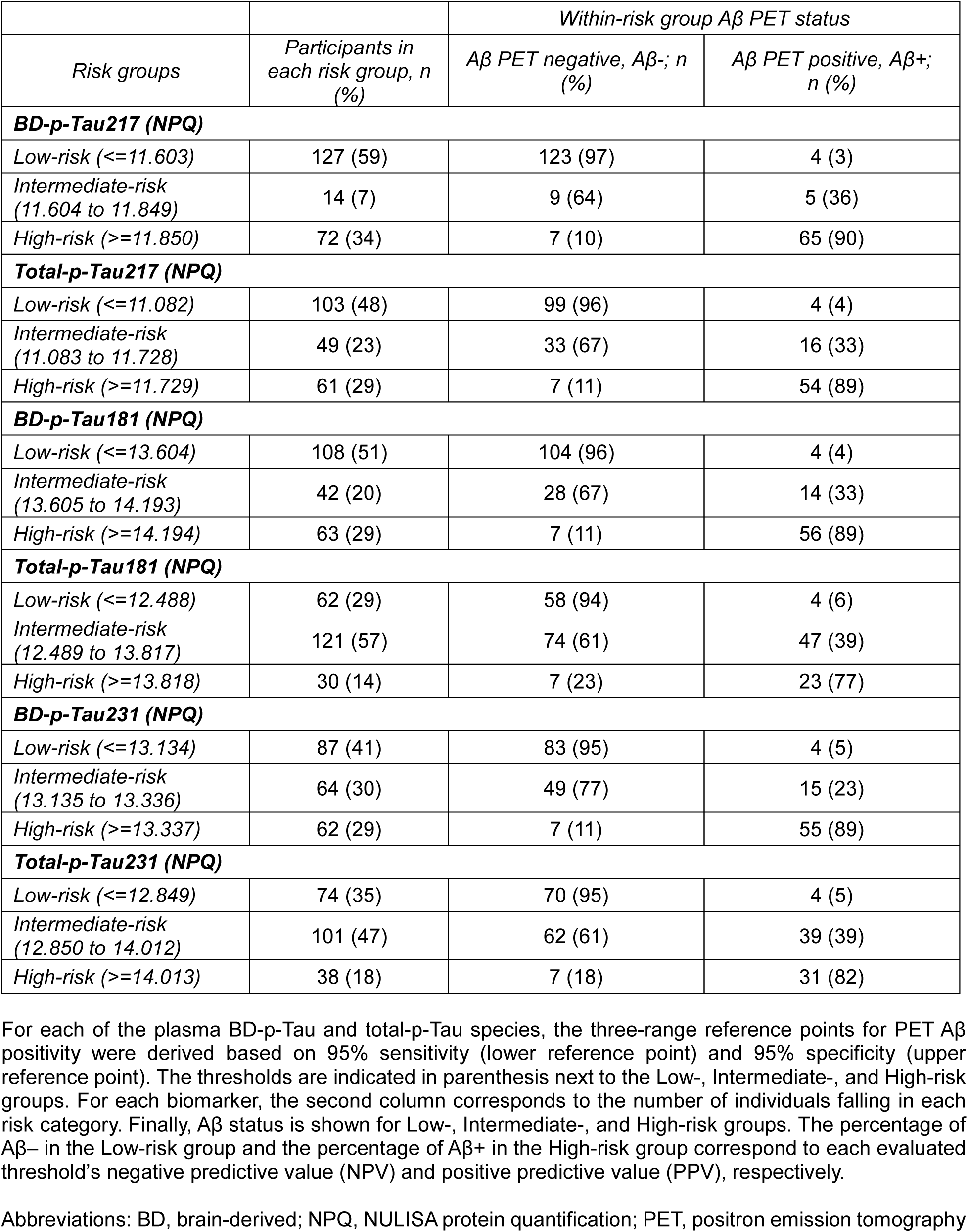
Three-range reference for Aβ-PET positivity.

|  |  | Within-risk group Aβ PET status |  |
| --- | --- | --- | --- |
| Risk groups | Participants in each risk group, n (%) | Aβ PET negative, Aβ-; n (%) | Aβ PET positive, Aβ+; n (%) |
| BD-p-Tau217 (NPQ) |  |  |  |
| Low-risk (<=11.603) | 127 (59) | 123 (97) | 4 (3) |
| Intermediate-risk (11.604 to 11.849) | 14 (7) | 9 (64) | 5 (36) |
| High-risk (>=11.850) | 72 (34) | 7 (10) | 65 (90) |
| Total-p-Tau217 (NPQ) |  |  |  |
| Low-risk (<=11.082) | 103 (48) | 99 (96) | 4 (4) |
| Intermediate-risk (11.083 to 11.728) | 49 (23) | 33 (67) | 16 (33) |
| High-risk (>=11.729) | 61 (29) | 7 (11) | 54 (89) |
| BD-p-Tau181 (NPQ) |  |  |  |
| Low-risk (<=13.604) | 108 (51) | 104 (96) | 4 (4) |
| Intermediate-risk (13.605 to 14.193) | 42 (20) | 28 (67) | 14 (33) |
| High-risk (>=14.194) | 63 (29) | 7 (11) | 56 (89) |
| Total-p-Tau181 (NPQ) |  |  |  |
| Low-risk (<=12.488) | 62 (29) | 58 (94) | 4 (6) |
| Intermediate-risk (12.489 to 13.817) | 121 (57) | 74 (61) | 47 (39) |
| High-risk (>=13.818) | 30 (14) | 7 (23) | 23 (77) |
| BD-p-Tau231 (NPQ) |  |  |  |
| Low-risk (<=13.134) | 87 (41) | 83 (95) | 4 (5) |
| Intermediate-risk (13.135 to 13.336) | 64 (30) | 49 (77) | 15 (23) |
| High-risk (>=13.337) | 62 (29) | 7 (11) | 55 (89) |
| Total-p-Tau231 (NPQ) |  |  |  |
| Low-risk (<=12.849) | 74 (35) | 70 (95) | 4 (5) |
| Intermediate-risk (12.850 to 14.012) | 101 (47) | 62 (61) | 39 (39) |
| High-risk (>=14.013) | 38 (18) | 7 (18) | 31 (82) |

For each of the plasma BD-p-Tau and total-p-Tau species, the three-range reference points for PET Aβ positivity were derived based on 95% sensitivity (lower reference point) and 95% specificity (upper reference point). The thresholds are indicated in parenthesis next to the Low-, Intermediate-, and High-risk groups. For each biomarker, the second column corresponds to the number of individuals falling in each risk category. Finally, Aβ status is shown for Low-, Intermediate-, and High-risk groups. The percentage of Aβ - in the Low-risk group and the percentage of Aβ + in the High-risk group correspond to each evaluated threshold's negative predictive value (NPV) and positive predictive value (PPV), respectively.
Abbreviations: BD, brain-derived; NPQ, NULISA protein quantification; PET, positron emission tomography

**Figure 2.**
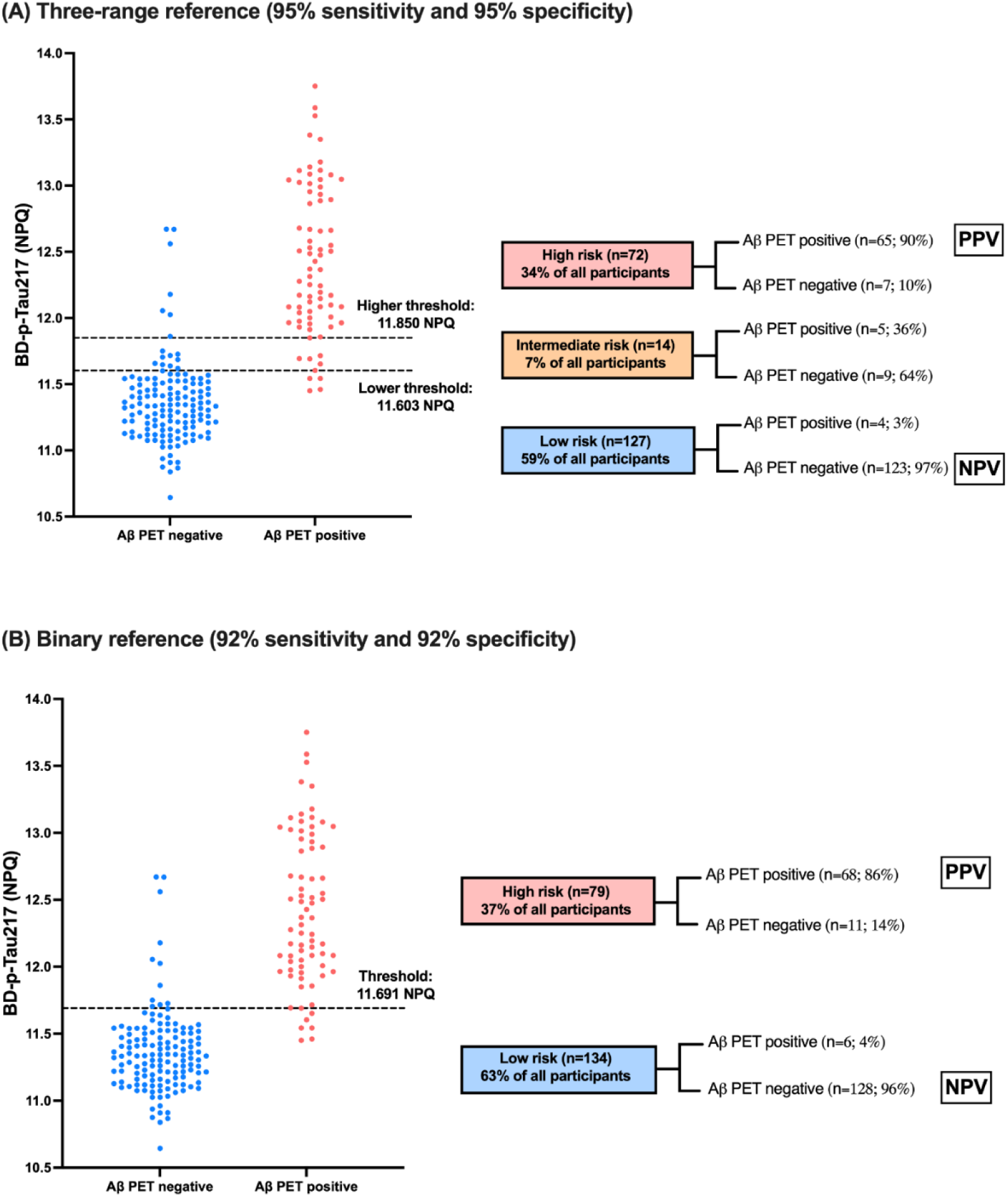
Three-range reference and binary reference for Aβ-PET positivity using plasma BD-p-Tau217. (A) Three-range reference and (B) binary reference for Aβ-PET positivity. Distribution of plasma BD-p-Tau217. The blue dots corresponded to individuals who are Aβ-PET negative and red dots to individuals who are Aβ-PET positive. In the three-range reference (A), the lower dashed line demonstrates where the 95% sensitivity Low-risk threshold falls on the distribution, with the upper line corresponding to the 95% specificity High-risk threshold. In the binary reference (B), the dashed line demonstrates the threshold derived by maximizing Youden index. On the right of each graph, the flowchart demonstrated the overall accuracy of each workflow. Abbreviations: BD, brain-derived; NPQ, NULISA protein quantification; NPV, negative predictive value; PET, positron emission tomography; PPV, positive predictive value

### 3.5 Binary reference of plasma BD- and total-p-Tau species for abnormal amyloid pathology

We next derived binary reference point for PET Aβ+ by maximizing the Youden index (**Supplementary Table S3** and **Figure 2B**). For plasma BD-p-Tau217, a cut-off point of 11.691 NPQ yielded the highest Youden index, achieving a sensitivity and specificity of 92%, as well as NPV and PPV of 96% and 86% respectively. No other plasma BD- and total-p-Tau species reached >90% in both sensitivity and specificity, although these p-Tau species achieved high NPVs between 89% and 95%, as well as PPVs between 73% and 83%, except for total-p-Tau181 which yielded a PPV of 57%.

### 3.6 Prognostic performance of the plasma BD-p-Tau217 reference ranges

We assessed the prognostic utility of the plasma BD-p-Tau217-derived three-range reference (**Figure 3**). Participants underwent annual cognitive assessments for up to 5 years (mean [SD] follow-up duration = 52 [13] months, relative to blood collection). As compared with the low-risk group (n=127), the high-risk group (n=72) demonstrated worse cognitive performance at baseline for MMSE (*β* = -3.74, *p* < 0.001) and CDR-SB (*β* = 2.17, *p* < 0.001). Longitudinally, cognitive performance decreased in both groups over time, with significantly higher rates in the high-risk group compared with the low-risk group (MMSE: *β* = -0.822, *p* < 0.001; CDR-SB: *β* = 0.799, *p* < 0.001). We also assessed the prognostic utility of the plasma BD-p-Tau217-derived binary reference (**Supplementary Figure S4**). Similar results were observed: as compared with the low-risk group (n=134), the high-risk group (n=79) had poorer baseline cognitive performance (MMSE: *β* = -3.29, *p* < 0.001; CDR-SB: *β* = 1.77, *p* < 0.001) and faster cognitive decline (MMSE: *β* = -0.742, *p* < 0.001; CDR-SB: *β* = 0.736, *p* < 0.001).

**Figure 3.**
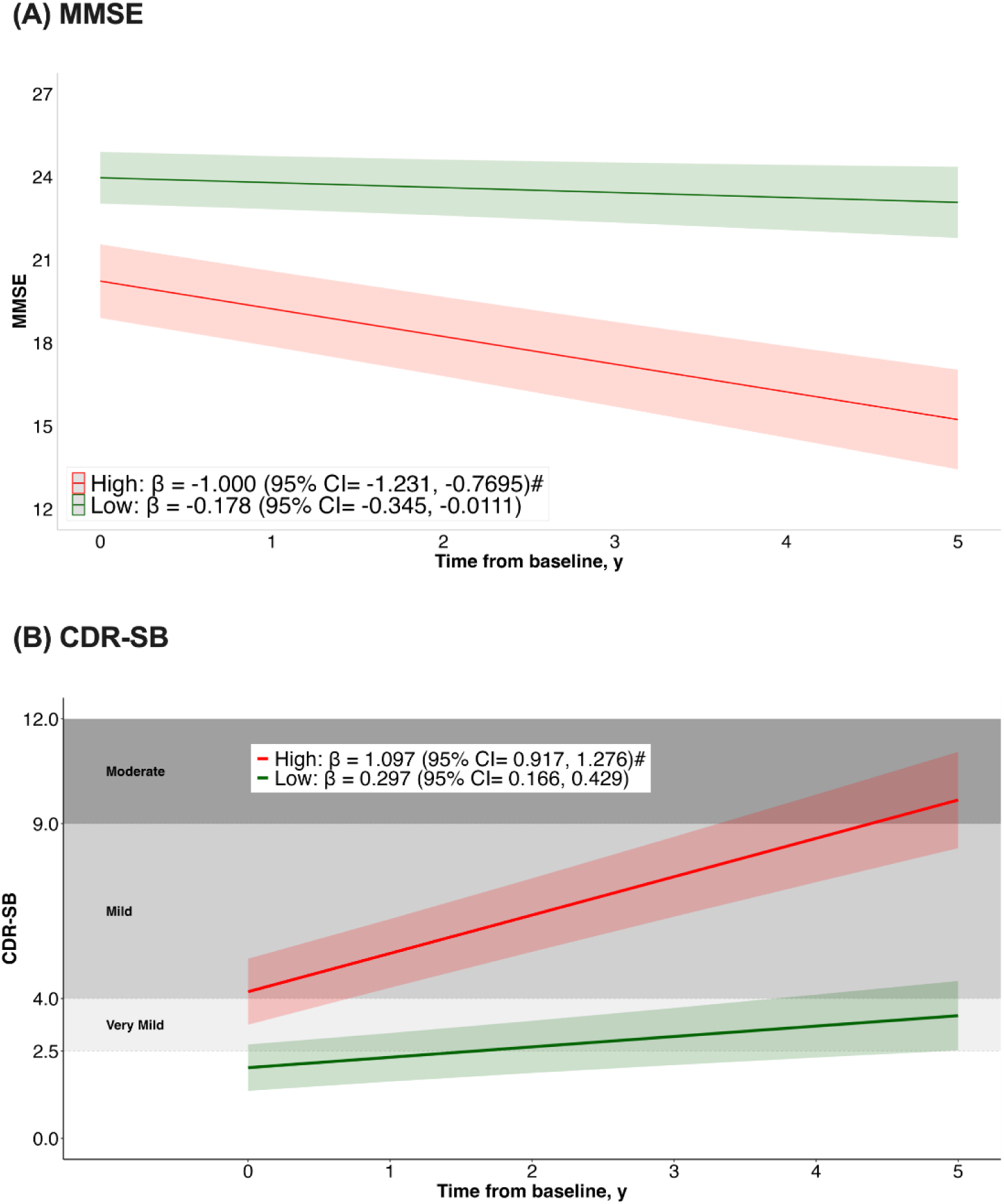
Prognostic performance of the plasma BD-p-Tau217-derived risk groups. Associations between plasma BD-p-Tau217-derived risk groups with baseline and longitudinal cognitive performance. Trajectory plots indicate the mean longitudinal trajectories (solid line) of (A) MMSE and (B) CDR-SB, and associated 95% confidence intervals (shaded areas), estimated with linear-mixed effects models. Trajectories are stratified based on the plasma BD-p-Tau217-derived risk groups for PET Aβ positivity (Low-risk [green] vs. High-risk [red]; derived from the three-range reference), modeled with an interaction between risk groups and time. Models included random slopes and intercepts and were adjusted for age, sex, and years of education. For CDR-SB, shaded background regions indicate dementia severity categories based on CDR-SB thresholds from [40]: 2.5–4.0 (very mild), 4.5–9.0 (mild), 9.5–15.5 (moderate), and 16.0–18.0 (severe). #Indicates longitudinal trajectories that are significantly different from Low-risk group. Abbreviations: BD, brain-derived; MMSE, Mini-Mental State Examination; CDR-SB, Clinical Dementia Rating Sum of Boxes; PET, positron emission tomography

Finally, we assessed the utility of the plasma BD-p-Tau217-derived three-range reference for predicting risk of progression to dementia (**Supplementary Table S4A and Supplementary Figure S5A**). Of the 151 non-dementia (CN and CIND) participants at baseline, 33 (22%) developed dementia (mean [SD] follow-up duration = 54 [11] months, relative to blood collection). 59% (23/39) of the high-risk group developed dementia compared to 8% (8/100) in the low-risk group, and 17% (2/12) in the intermediate-risk group. Cox regression (**Supplementary Table S4A**) showed an increased risk of progression to dementia in the high-risk group (hazard ratio [HR] = 5.62, 95% CI = 2.39 to 13.2, *p* < 0.001) compared to the low-risk group. **Supplementary Figure S5A** depicts the Kaplan-Meier survival curve. We also evaluated the utility of the plasma BD-p-Tau217-derived binary reference (**Supplementary Table S4B and Supplementary Figure S5B**). 53% (24/45) of the high-risk group developed dementia compared to 8% (9/106) in the low-risk group. Cox regression (**Supplementary Table S4B**) showed an increased risk of progression to dementia in the high-risk group (hazard ratio [HR] = 5.29, 95% CI = 2.38 to 11.8, *p* < 0.001). **Supplementary Figure S5B** depicts the Kaplan-Meier survival curve.

## DISCUSSION

In this study of an Asian-based cohort of older people known to have high baseline CeVD burden [23], we present evidence that a novel biomarker, plasma brain-derived (BD)-p-Tau217 outperformed total-p-Tau217 as well as the BD- and total-p-Tau species of Tau181 and Tau231 in detecting brain amyloid pathology, with AUC = 0.965. Furthermore, individuals in the plasma BD-p-Tau217-derived high-risk group exhibited faster, longitudinally-assessed cognitive decline.

With the recent approval of two blood p-Tau tests by the U.S. Food and Drug Administration (FDA) to aid in the diagnosis of AD [31], plasma p-Tau biomarkers with high diagnostic accuracy for detecting amyloid pathology have the potential to become accessible, cost-effective tools for AD evaluation in clinical settings. Among the plasma p-Tau species studied to date, p-Tau217 has emerged as the best-performing biomarker for detecting abnormal brain amyloid burden, with comparable performance across different assay platforms [1, 2]. However, these widely used assays do not differentiate between brain-derived LMW Tau versus HMW Tau from PNS and other organs. In the case of studies on ALS highlighted in the introduction [20, 21], measurements of both BD-and total p-Tau217 might have shown differential alterations in the biomarkers, thus providing potentially useful information on stage / severity of degeneration of motor cortex and brainstem motor neurons as well as denervation at the neuromuscular junction. In the case of AD, the use of BD-p-Tau217 putatively avoids the detection of p-Tau species secreted by peripheral organs (up to 80% of total Tau [32, 33], also see **Figure 1C**) which are not reflective of Tau status in the brain, thus enabling improved utility as the current study has shown. Indeed, the outstanding diagnostic performance of plasma BD-p-Tau217 in detecting abnormal brain amyloid burden was consistent in the main cohort (**Table 2**, n=213) as well as in sensitivity analyses of smaller subsets (**Supplementary Table S1**, n=114; **Supplementary Table S2**, n=152). Notably, plasma BD-p-Tau217 was significantly better than total-p-Tau217 and other BD- and total-p-Tau species across all analyses, indicating the robustness of our findings (**Figure 1D**).

This study also aimed to establish reference points for the NULISA plasma p-Tau markers based on abnormal amyloid pathology. We first evaluated a three-range approach using a threshold of 95% sensitivity and specificity, which is more stringent than the 90% sensitivity and specificity currently recommended by the Global CEO Initiative (GCI) on Alzheimer’s disease for blood-based biomarkers to be used as diagnostic tests for amyloid pathology [30]. We observed that all p-Tau biomarkers achieved largely similar NPVs (94% to 97%) and PPVs (77% to 90%), with plasma BD-p-Tau217 showing the highest NPV and PPV of 97% and 90%, respectively. When comparing the proportion of participants classified based on risk of amyloid positivity, plasma BD-p-Tau217 resulted in only 7% of individuals determined as having intermediate risk which necessitates further confirmatory testing with CSF markers or amyloid PET, whereas other plasma BD-p-Tau and total-p-Tau species yielded at least 20% to 57% of participants in the intermediate risk zone. Since intermediate results are not informative of brain amyloid status, the GCI Workgroup recommended that a blood test should have intermediate values in no more than 15% of the cohort [30], a benchmark met only by BD-p-Tau217 (**Figure 1E** and **Figure 2**).

The three-range threshold approach was introduced to enhance the overall performance of plasma biomarkers for identifying amyloid burden - however, this approach is not required if a single cut-off yields acceptable accuracy [30]. Using the conventional Youden index threshold for binary classification, plasma BD-p-Tau217 was the only biomarker with both sensitivity and specificity above 90% (specifically 92%), thus meeting the GCI’s recommended minimum benchmark of 90%. In contrast, other plasma p-Tau species, including total-p-Tau217 measured by NULISA or Simoa approached this benchmark but did not meet it, consistent with previous findings [1, 12]. Additionally, the binary plasma BD-p-Tau217 threshold yielded high NPV and PPV of 96% and 86%, respectively. Taken together, our data suggest that of the six biomarkers we evaluated in the current study, plasma BD-p-Tau217 stood out as the only biomarker that achieved all the CGI Workgroup’s proposed benchmarks for both the three-range and single cut-off diagnostic performance for brain amyloid pathology.

In this study, we further assessed the prognostic utility of plasma BD-p-Tau217-derived risk groups in predicting cognitive decline. Compared with the low-risk group, participants in the high-risk group, defined by either the three-range or binary approach, exhibited faster rates of cognitive decline (**Figure 1F** and **Figure 3**), as well as higher risk of progression to dementia (**Supplementary Table S4 and Supplementary Figure S5).** This aligns with the association between AD pathology and subsequent neurodegeneration and cognitive decline [5]. While AD is the most common cause of cognitive impairment and dementia, co-pathologies such as cerebrovascular disease, alpha-synucleinopathy, TAR DNA-binding protein 43 (TDP-43) inclusions, and neuroinflammation are frequently observed in AD patients [34, 35]. These co-pathologies may exacerbate disease trajectory, leading to more rapid cognitive decline [5, 34–36]. In this context, it has recently been proposed that a multimodal biomarker profile that more comprehensively reflects diverse neuropathophysiological processes may enhance risk stratification and prognostication [5]. Future studies should therefore evaluate combinations of plasma BD-p-Tau217 with biomarkers of non-AD pathologies, as well as key pathophysiological processes, for predicting cognitive decline.

Several limitations were apparent in our study. First, the cohort is based in Asia, and further validation of current findings in larger, more diverse cohorts that more closely reflect the intended-use populations is warranted, since variable demographic factors, risk factors and comorbidities may influence the clinical performance and applicability of blood biomarkers in specific populations [37]. Second, beyond amyloid PET, future studies should examine the utility of plasma BD-p-Tau217 in detecting Tau PET positivity. There is also growing interest in using plasma biomarkers, including plasma p-Tau217, to distinguish between the biological PET stages of AD [11, 38], as proposed by the Revised Criteria for Diagnosis and Staging of Alzheimer’s Disease [5]. Given the superior diagnostic performance of plasma BD-p-Tau217, it would be worthwhile to evaluate the performance and thresholds of this biomarker for predicting biological PET staging. Third, we recognize the variability in the time interval between blood collection and amyloid PET among participants, where blood collection occurred on or before PET imaging in the majority of the participants (specifically, in 203 (95%) participants). However, given the progressive and relatively stable nature of amyloid PET findings, participants classified as PET Aβ-at the later imaging time point would reasonably be expected to have plasma biomarker results reflective of Aβ-from the earlier blood draw. Given the slow rate of brain amyloid accumulation [39], it is also plausible that participants classified as PET Aβ+ at the time of imaging would have been PET Aβ+ at the earlier blood collection. As a sensitivity analysis, we have confirmed that the AUC analyses in participants with amyloid PET within three years of blood collection (n=114, **Supplementary Table S1**) corroborated the main findings that plasma BD-p-Tau217 outperformed other BD- and total-p-Tau species. Nevertheless, follow-up studies with shorter time difference between blood collection and PET imaging are recommended to validate these findings.

In conclusion, the findings of this study highlight the potential clinical utility of plasma BD-p-Tau217 for both diagnosis of amyloid pathology and prognosis of AD-associated cognitive impairments. Our results support rigorous validation of this biomarker in larger, more ethnically diverse cohorts in the backdrop of imminent adoption of blood biomarkers as a routine workup for AD-associated cognitive decline and dementia.

## ACKNOWLEDGMENTS

We are grateful to the patients and their families for their participation in this study. We acknowledge the coordinator and rater teams from the Memory, Ageing and Cognition Centre for assistance with participant recruitment and assessment. This study was supported by Alamar Biosciences’ Technology Access Program, which played no role in the writing and editing of this manuscript, or the decision to publish.

## CONFLICT OF INTEREST STATEMENT

Author disclosures are available in the supporting information of the finalized version of manuscript.

## FUNDING

National Medical Research Council of Singapore Grant/Award Number: MOH-000707-01; Singapore Ministry of Education Academic Research Fund Award Number: NUHSRO/2024/028/T1/Seed-Sep23/06.

## CONSENT STATEMENT

Approval for the study was obtained from the Singapore National Healthcare Group Domain-Specific Review Board (2018/00996, 2015/00406, and 2015/00441). Written informed consent was obtained from study participants or their next of kin.

## DATA AVAILABILITY STATEMENT

Anonymized datasets generated during and/or analyzed in the current study are available from the corresponding author upon reasonable request.

## AUTHOR CONTRIBUTIONS

Mitchell K. P. Lai had full access to all data in the study and takes responsibility for the integrity of the data and the accuracy of the data analysis. Study concept and design: Joyce R. Chong and Mitchell K. P. Lai. Acquisition, analysis, or interpretation of data: Joyce R. Chong, Saima Hilal, Narayanaswamy Venketasubramanian, Michael Schöll, Nicholas J. Ashton, Henrik Zetterberg, Christopher P. Chen, Mitchell K. P. Lai. Drafting of the manuscript: Joyce R. Chong and Mitchell K. P. Lai. Critical revision of the manuscript for important intellectual content: Joyce R. Chong, Saima Hilal, Narayanaswamy Venketasubramanian, Michael Schöll, Nicholas J. Ashton, Henrik Zetterberg, Christopher P. Chen, Mitchell K. P. Lai. Administrative, technical, or material support: Joyce R. Chong. Obtained funding: Christopher P. Chen and Mitchell K. P. Lai. Supervision: Christopher P. Chen and Mitchell K. P. Lai.

## Notes

### Competing Interest Statement

The authors have declared no competing interest.

### Funding Statement

This study was funded by the National Medical Research Council of Singapore Grant/Award Number: MOH-000707-01; Singapore Ministry of Education Academic Research Fund Award Number: NUHSRO/2024/028/T1/Seed-Sep23/06.

### Author Declarations

Ethics committee/IRB of Singapore National Healthcare Group gave ethical approval for this work.

### Summary of Updates

1. Added new Supplementary Table S4 with associated links and description in manuscript. 2. Added new Supplementary Figure S2 with associated links and description in manuscript. 3. Modified Supplementary Figure S4B with associated links and description in manuscript. 4. Added new Supplementary Figure S5 with associated links and description in manuscript. 5. Modified Figure 1 with associated links and description in manuscript.

## REFERENCES

1. Therriault, J., et al., Blood phosphorylated tau for the diagnosis of Alzheimer’s disease: a systematic review and meta-analysis. Lancet Neurol, 2025. 24(9): p. 740–752.

2. Ashton, N.J., et al., The Alzheimer’s Association Global Biomarker Standardization Consortium (GBSC) plasma phospho-tau Round Robin study. Alzheimers Dement, 2025. 21(2): p. e14508.

3. Warmenhoven, N., et al., A comprehensive head-to-head comparison of key plasma phosphorylated tau 217 biomarker tests. Brain, 2025. 148(2): p. 416–431.

4. Schindler, S.E., et al., Head-to-head comparison of leading blood tests for Alzheimer’s disease pathology. medRxiv, 2024.

5. Jack, C.R., Jr., et al., Revised criteria for diagnosis and staging of Alzheimer’s disease: Alzheimer’s Association Workgroup. Alzheimers Dement, 2024. 20(8): p. 5143–5169.

6. Ashton, N.J., et al., Diagnostic Accuracy of a Plasma Phosphorylated Tau 217 Immunoassay for Alzheimer Disease Pathology. JAMA Neurol, 2024. 81(3): p. 255–263.

7. Rea Reyes, R.E., et al., Targeted proteomic biomarker profiling using NULISA in a cohort enriched with risk for Alzheimer’s disease and related dementias. Alzheimers Dement, 2025. 21(5): p. e70166.

8. Therriault, J., et al., Comparison of two plasma p-tau217 assays to detect and monitor Alzheimer’s pathology. EBioMedicine, 2024. 102: p. 105046.

9. Palmqvist, S., et al., Plasma phospho-tau217 for Alzheimer’s disease diagnosis in primary and secondary care using a fully automated platform. Nat Med, 2025. 31(6): p. 2036–2043.

10. Zeng, X., et al., Multi-analyte proteomic analysis identifies blood-based neuroinflammation, cerebrovascular and synaptic biomarkers in preclinical Alzheimer’s disease. Molecular Neurodegeneration, 2024. 19(1): p. 68.

11. Feizpour, A., et al., Detection and staging of Alzheimer’s disease by plasma pTau217 on a high throughput immunoassay platform. EBioMedicine, 2024. 109: p. 105405.

12. Chong, J.R., et al., Clinical utility of plasma p-tau217 in identifying abnormal brain amyloid burden in an Asian cohort with high prevalence of concomitant cerebrovascular disease. Alzheimers Dement, 2025. 21(2): p. e14502.

13. Huang, K.L., et al., The Taiwan-ADNI workflow toward integrating plasma p-tau217 into prediction models for the risk of Alzheimer’s disease and tau burden. Alzheimers Dement, 2025. 21(1): p. e14297.

14. Kwon, H.S., et al., Comparison of plasma p-tau217 and p-tau181 in predicting amyloid positivity and prognosis among Korean memory clinic patients. Scientific Reports, 2025. 15(1): p. 7791.

15. Lin, Y.S., et al., Cross-cultural validation of plasma p-tau217 and p-tau181 as precision biomarkers for amyloid PET positivity: An East Asian study in Taiwan and Korea. Alzheimers Dement, 2025. 21(1): p. e14565.

16. Wang, J., et al., Diagnostic accuracy of plasma p-tau217/Aβ42 for Alzheimer’s disease in clinical and community cohorts. Alzheimers Dement, 2025. 21(3): p. e70038.

17. Janelidze, S., et al., A comparison of p-tau assays for the specificity to detect tau changes in Alzheimer’s disease. Alzheimers Dement, 2025. 21(4): p. e70208.

18. Donison, N., et al., Cellular and molecular mechanisms of pathological tau phosphorylation in traumatic brain injury: implications for chronic traumatic encephalopathy. Mol Neurodegener, 2025. 20(1): p. 56.

19. Buchholz, S. and H. Zempel, The six brain-specific TAU isoforms and their role in Alzheimer’s disease and related neurodegenerative dementia syndromes. Alzheimers Dement, 2024. 20(5): p. 3606–3628.

20. Abu-Rumeileh, S., et al., Phosphorylated tau 181 and 217 are elevated in serum and muscle of patients with amyotrophic lateral sclerosis. Nat Commun, 2025. 16(1): p. 2019.

21. Vacchiano, V., et al., Elevated plasma p-tau181 levels unrelated to Alzheimer’s disease pathology in amyotrophic lateral sclerosis. J Neurol Neurosurg Psychiatry, 2023. 94(6): p. 428–435.

22. Lim, M.J., et al., HARMONISATION - A multimodal prospective study of vascular cognitive impairment in multi-ethnic Asians: Cohort profile, progress, current contributions, and future impact. J Alzheimers Dis, 2025. 108(4): p. 1452–1474.

23. Chong, J.R., et al., Plasma P-tau181 to Aβ42 ratio is associated with brain amyloid burden and hippocampal atrophy in an Asian cohort of Alzheimer’s disease patients with concomitant cerebrovascular disease. Alzheimers Dement, 2021. 17(10): p. 1649–1662.

24. Chong, J.R., et al., Brain atrophy and white matter hyperintensities are independently associated with plasma neurofilament light chain in an Asian cohort of cognitively impaired patients with concomitant cerebral small vessel disease. Alzheimers Dement (Amst), 2023. 15(1): p. e12396.

25. Chong, J.R., et al., Association of plasma GFAP with elevated brain amyloid is dependent on severity of white matter lesions in an Asian cognitively impaired cohort. Alzheimers Dement (Amst), 2024. 16(2): p. e12576.

26. Saridin, F.N., et al., Cerebrovascular disease in suspected non-Alzheimer’s pathophysiology and cognitive decline over time. Eur J Neurol, 2022. 29(7): p. 1922–1929.

27. Feng, W., et al., NULISA: a proteomic liquid biopsy platform with attomolar sensitivity and high multiplexing. Nature Communications, 2023. 14(1): p. 7238.

28. Robin, X., et al., pROC: an open-source package for R and S+ to analyze and compare ROC curves. BMC Bioinformatics, 2011. 12: p. 77.

29. Brum, W.S., et al., A two-step workflow based on plasma p-tau217 to screen for amyloid β positivity with further confirmatory testing only in uncertain cases. Nat Aging, 2023. 3(9): p. 1079–1090.

30. Schindler, S.E., et al., Acceptable performance of blood biomarker tests of amyloid pathology - recommendations from the Global CEO Initiative on Alzheimer’s Disease. Nat Rev Neurol, 2024. 20(7): p. 426–439.

31. Kavanagh, K., Blood tests are now approved for Alzheimer’s: how accurate are they? Nature, 2025. 646(8087): p. 1031–1032.

32. Ma, X.J., et al. Highly sensitive multiplexed detection of brain-derived Tau and phosphorylated Tau proteoforms with the NULISA technology. in Alzheimer’s Association International Conference. 2025. Toronto.

33. Barthélemy, N.R., et al., Blood plasma phosphorylated-tau isoforms track CNS change in Alzheimer’s disease. J Exp Med, 2020. 217(11).

34. Rabinovici, G.D., et al., Multiple comorbid neuropathologies in the setting of Alzheimer’s disease neuropathology and implications for drug development. Alzheimers Dement (N Y), 2017. 3(1): p. 83–91.

35. Tosun, D., et al., Identifying individuals with non-Alzheimer’s disease co-pathologies: A precision medicine approach to clinical trials in sporadic Alzheimer’s disease. Alzheimers Dement, 2024. 20(1): p. 421–436.

36. Collij, L.E., et al., Lewy body pathology exacerbates brain hypometabolism and cognitive decline in Alzheimer’s disease. Nature Communications, 2024. 15(1): p. 8061.

37. Schöll, M., et al., Challenges in the practical implementation of blood biomarkers for Alzheimer’s disease. Lancet Healthy Longev, 2024. 5(10): p. 100630.

38. Doecke, J.D., et al., Multivariate models using NULISAseq and plasma p-tau217 for staging Alzheimer’s disease. Alzheimers Dement, 2025. 21(10): p. e70793.

39. Villemagne, V.L., et al., Amyloid β deposition, neurodegeneration, and cognitive decline in sporadic Alzheimer’s disease: a prospective cohort study. Lancet Neurol, 2013. 12(4): p. 357–67.

40. O’Bryant, S.E., et al., Validation of the new interpretive guidelines for the clinical dementia rating scale sum of boxes score in the national Alzheimer’s coordinating center database. Arch Neurol, 2010. 67(6): p. 746–9.

