## Supplementary for "Plasma brain-derived p-Tau217 outperforms other p-Tau species in detecting abnormal brain amyloid in an Asian cohort of older people with cerebrovascular disease burden"

### Supplementary Materials

#### Supplementary Table S1. Diagnostic performance of the brain-derived and total p-Tau species in identifying abnormal brain amyloid burden in participants with amyloid PET within three years of blood collection

| *PET Aβ+ = 45, PET Aβ- = 69* | *AUC (95% CI)* | *p-value when compared to BD-p-Tau217* |
|---|---|---|
| *BD-p-Tau217* | 0.947 (0.908, 0.985) | NA |
| *Total-p-Tau217* | 0.920 (0.871, 0.970) | **0.032** |
| *BD-p-Tau181* | 0.912 (0.858, 0.966) | **0.020** |
| *BD-p-Tau231* | 0.912 (0.858, 0.967) | **0.048** |
| *Total-p-Tau231* | 0.846 (0.773, 0.920) | **<0.001** |
| *Total-p-Tau181* | 0.789 (0.706, 0.871) | **<0.001** |

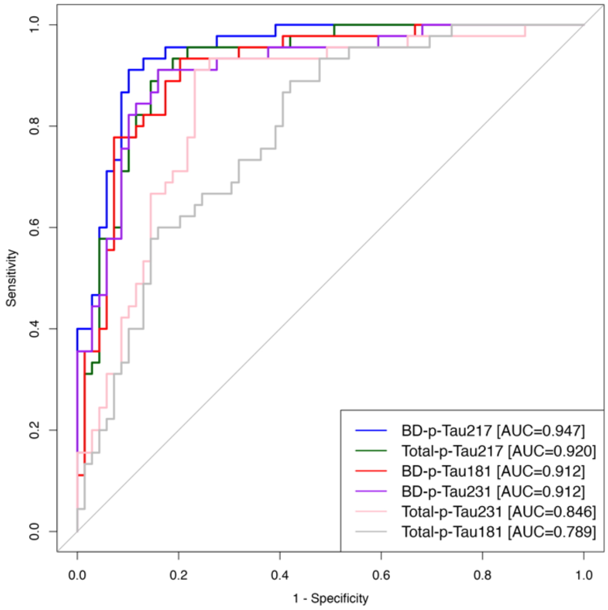


AUC and the associated 95% CI for each of the plasma BD-p-Tau (BD-p-Tau217, BD-p-Tau181, BD-p-Tau231) and total-p-Tau (total-p-Tau217, total-p-Tau181, total-p-Tau231) species, in predicting Aβ-PET positivity. Plasma BD-p-Tau217 outperformed other BD-p-Tau and total-p-Tau species in detecting abnormal brain amyloid burden ($p \leq 0.048$). P-values (**bold** fonts denoting statistical significance) for pairwise comparisons of AUCs were derived from DeLong tests.

Abbreviations: AUC, area under the receiver-operating characteristic (ROC) curve; BD, brain-derived; CI, confidence interval; PET, positron emission tomography

**Supplementary Table S2. Comparisons of NULISA brain-derived and total p-Tau species with Simoa p-Tau217 in identifying abnormal brain amyloid burden**

| *PET Aβ+ = 52, PET Aβ- = 100* | *AUC (95% CI)* | *p-value when compared to BD-p-Tau217* |
|---|---|---|
| *BD-p-Tau217* | 0.974 (0.953, 0.996) | NA |
| *Total-p-Tau217* | 0.947 (0.908, 0.985) | **0.015** |
| *BD-p-Tau181* | 0.943 (0.905, 0.980) | **0.010** |
| *BD-p-Tau231* | 0.938 (0.897, 0.978) | **0.027** |
| *Simoa p-Tau217* | 0.917 (0.864, 0.970) | **0.007** |
| *Total-p-Tau231* | 0.881 (0.823, 0.939) | **<0.001** |
| *Total-p-Tau181* | 0.833 (0.767, 0.899) | **<0.001** |

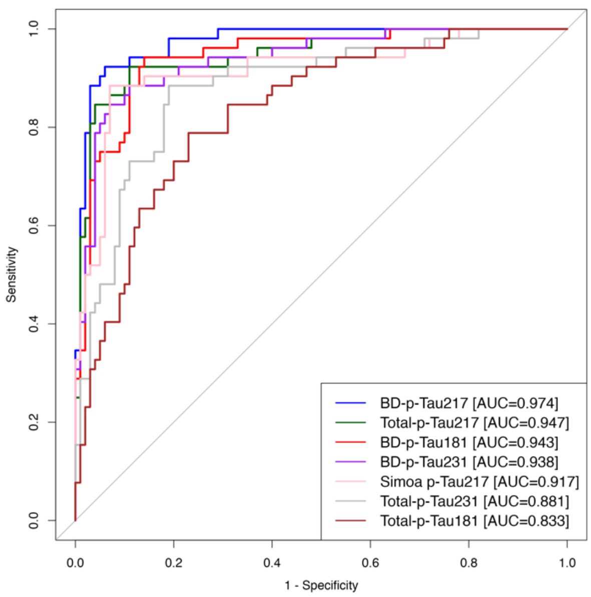


AUC and the associated 95% CI for each of the plasma BD-p-Tau (BD-p-Tau217, BD-p-Tau181, BD-p-Tau231) and total-p-Tau (total-p-Tau217, total-p-Tau181, total-p-Tau231) species, as well as Simoa p-Tau217, in predicting Aβ-PET positivity. Plasma BD-p-Tau217 outperformed other BD-p-Tau and total-p-Tau species, as well as Simoa p-Tau217 ($p = 0.007$), in detecting abnormal brain amyloid burden. The mean (SD) interval between blood collection and PET imaging was 39 (23) months. P-values (**bold** fonts denoting statistical significance) for pairwise comparisons of AUCs were derived from DeLong tests.

Abbreviations: AUC, area under the receiver-operating characteristic (ROC) curve; BD, brain-derived; CI, confidence interval; PET, positron emission tomography

### Supplementary Table S3. Binary reference for Aβ-PET positivity

| | | | | *Within-risk group Aβ PET status* | |
|---|---|---|---|---|---|
| *Risk groups* | *Specificity (%)* | *Sensitivity (%)* | *Participants in each risk group (n (%))* | *Aβ PET negative (Aβ-; n (%))* | *Aβ PET positive (Aβ+; n (%))* |
| ***BD-p-Tau217 (NPQ)*** | | | | | |
| *Low-risk (<11.691)* | 92.1 | 91.9 | 134 (63) | 128 (96) | 6 (4) |
| *High-risk (>=11.691)* | | | 79 (37) | 11 (14) | 68 (86) |
| ***Total-p-Tau217 (NPQ)*** | | | | | |
| *Low-risk (<11.541)* | 90.6 | 87.8 | 135 (63) | 126 (93) | 9 (7) |
| *High-risk (>=11.541)* | | | 78 (37) | 13 (17) | 65 (83) |
| ***BD-p-Tau181 (NPQ)*** | | | | | |
| *Low-risk (<13.825)* | 87.1 | 91.9 | 127 (60) | 121 (95) | 6 (5) |
| *High-risk (>=13.825)* | | | 86 (40) | 18 (21) | 68 (79) |
| ***Total-p-Tau181 (NPQ)*** | | | | | |
| *Low-risk (<12.901)* | 66.2 | 85.1 | 103 (48) | 92 (89) | 11 (11) |
| *High-risk (>=12.901)* | | | 110 (52) | 47 (43) | 63 (57) |
| ***BD-p-Tau231 (NPQ)*** | | | | | |
| *Low-risk (<13.259)* | 88.5 | 89.2 | 131 (62) | 123 (94) | 8 (6) |
| *High-risk (>=13.259)* | | | 82 (38) | 16 (20) | 66 (80) |
| ***Total-p-Tau231 (NPQ)*** | | | | | |
| *Low-risk (<13.270)* | 82.7 | 86.5 | 125 (59) | 115 (92) | 10 (8) |
| *High-risk (>=13.270)* | | | 88 (41) | 24 (27) | 64 (73) |

For each of the plasma BD-p-Tau and total-p-Tau species, the binary reference point for PET Aβ positivity was determined by maximizing the Youden index. The thresholds are indicated in parenthesis next to the Low- and High-risk groups. For each biomarker, the second and third column correspond to the specificity and sensitivity respectively, while the fourth column corresponds to the number of individuals falling in each risk category. Lastly, Aβ status is shown for Low- and High-risk groups. The percentage of Aβ- in the Low-risk group and the percentage of Aβ+ in the High-risk group correspond to each evaluated threshold's negative predictive value (NPV) and positive predictive value (PPV), respectively.

Abbreviations: BD, brain-derived; NPQ, NULISA protein quantification units; PET, positron emission tomography

**Supplementary Table S4. Cox regression analyses of risk for progression to dementia**

**(A) Plasma BD-p-Tau217-derived risk groups based on 3-range reference**

| | *Progression to dementia (Outcome)* | | | | |
|---|---|---|---|---|---|
| *Risk of amyloid PET positivity* | *Stable, n (%)* | *Declined, n (%)* | *Hazard ratio* | *95% CI* | *p-value* |
| Low-risk (n=100) | 92 (92) | 8 (8) | | Reference | |
| High-risk (n=39) | 16 (41) | 23 (59) | **5.62** | **(2.39, 13.2)** | **<0.001** |

**(B) Plasma BD-p-Tau217-derived risk groups based on binary reference**

| | *Progression to dementia (Outcome)* | | | | |
|---|---|---|---|---|---|
| *Risk of amyloid PET positivity* | *Stable, n (%)* | *Declined, n (%)* | *Hazard ratio* | *95% CI* | *p-value* |
| Low-risk (n=106) | 97 (92) | 9 (8) | | Reference | |
| High-risk (n=45) | 21 (47) | 24 (53) | **5.29** | **(2.38, 11.8)** | **<0.001** |

Cox regression of progression to dementia in follow-up among non-dementia participants (CN + CIND). Participants were stratified by risk of amyloid PET positivity (Low risk versus High risk), derived based on (A) the 3-range reference or (B) the binary reference of plasma BD-p-Tau217. Cox regression model adjusted for age, sex and education. P-values in **bold** fonts denote statistical significance.

Abbreviations: BD, brain-derived; CI, confidence interval; CIND, cognitive impairment no dementia; CN, cognitively normal; PET, positron emission tomography

### Supplementary Figure S1. Correlations between plasma brain-derived and total p-Tau species with PiB-PET SUVr

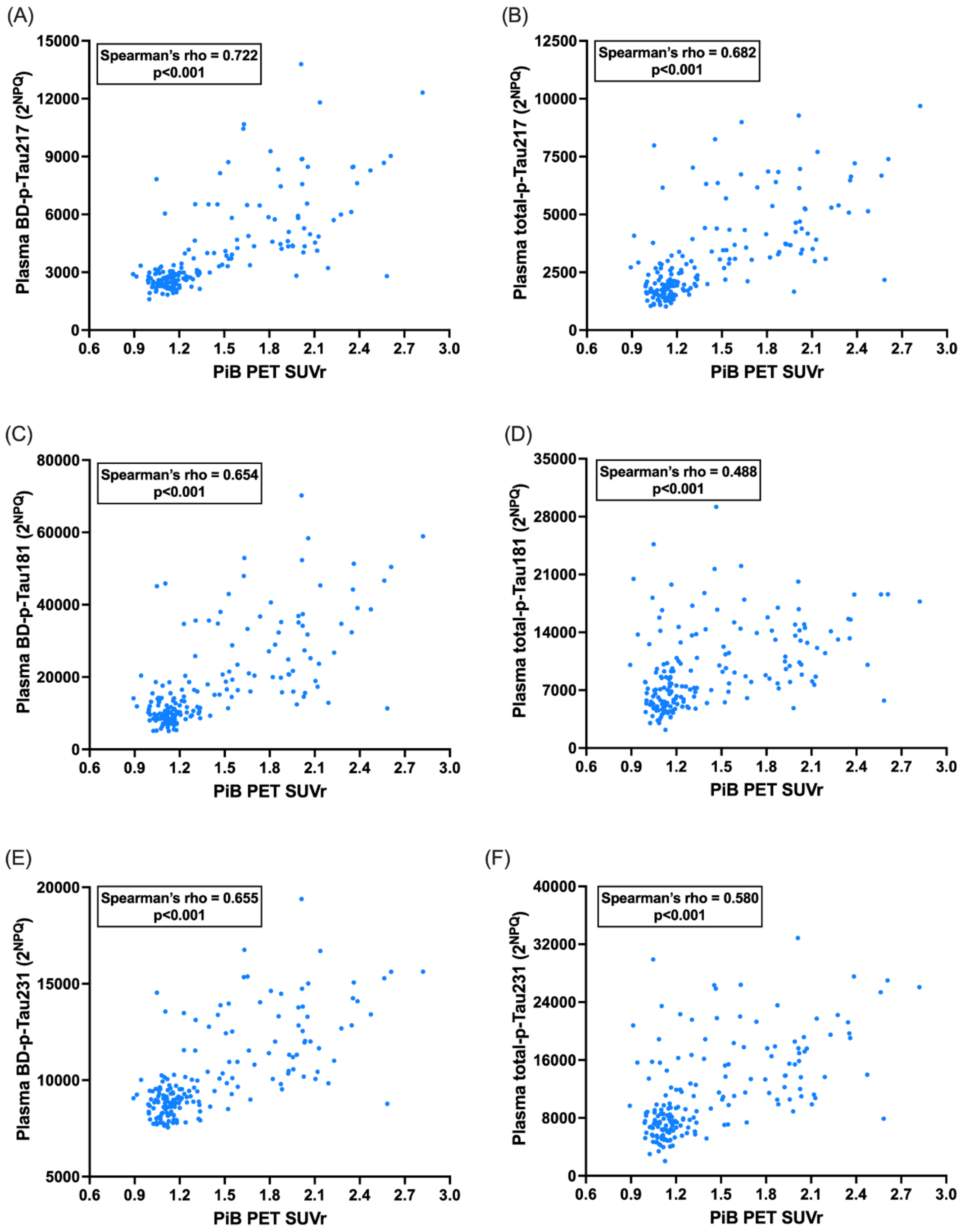


Correlations between each plasma BD-p-Tau (A, C and E) and total-p-Tau (B, D, F) species with PiB-PET SUVr.

Abbreviations: BD, brain-derived; PET, positron emission tomography; PiB, Pittsburgh compound B; SUVr, standardized uptake value ratio

### Supplementary Figure S2. Plasma brain-derived and total p-Tau species by clinical diagnosis and Aβ-PET status

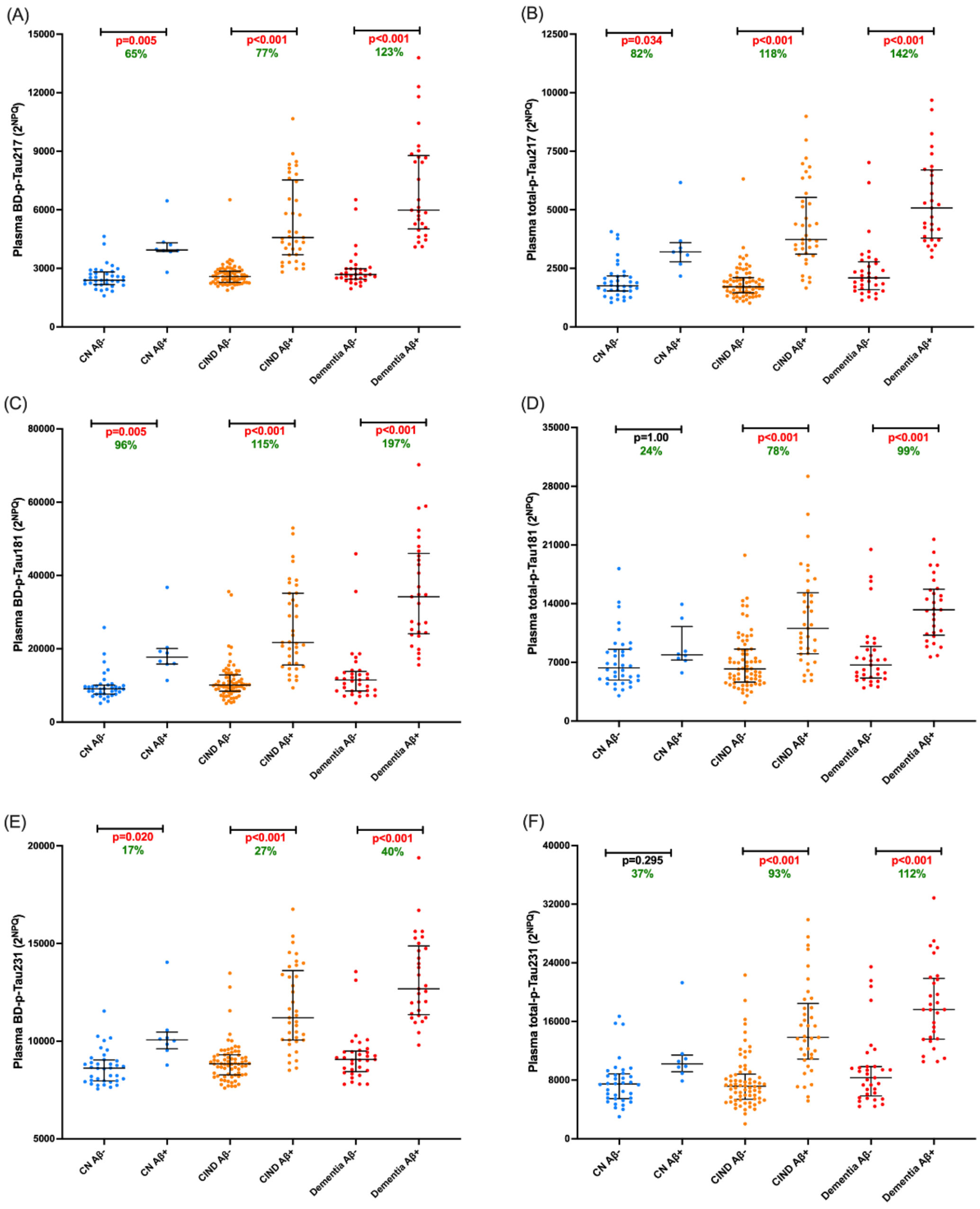


Plasma BD-p-Tau (A, C and E) and total-p-Tau (B, D, F) species by clinical diagnosis and Aβ-PET status. The graphs show the median and interquartile range. P-values derived from Kruskal–Wallis tests followed by *post-hoc* Dunn's tests with Bonferroni correction. Within each clinical diagnostic group, the percentage increase of plasma biomarker in the amyloid-positive (Aβ+) group from the amyloid-negative (Aβ–) group is shown in green (percentage increase = [(Median Aβ+ - Median Aβ-) / Median Aβ-] × 100%). Red fonts indicate significant p-values ($p < 0.05$).

Abbreviations: CN, cognitively normal; CIND, cognitive impairment no dementia; PET, positron emission tomography

### Supplementary Figure S3. Three-range reference for PET amyloid positivity using NULISA plasma total-p-Tau217

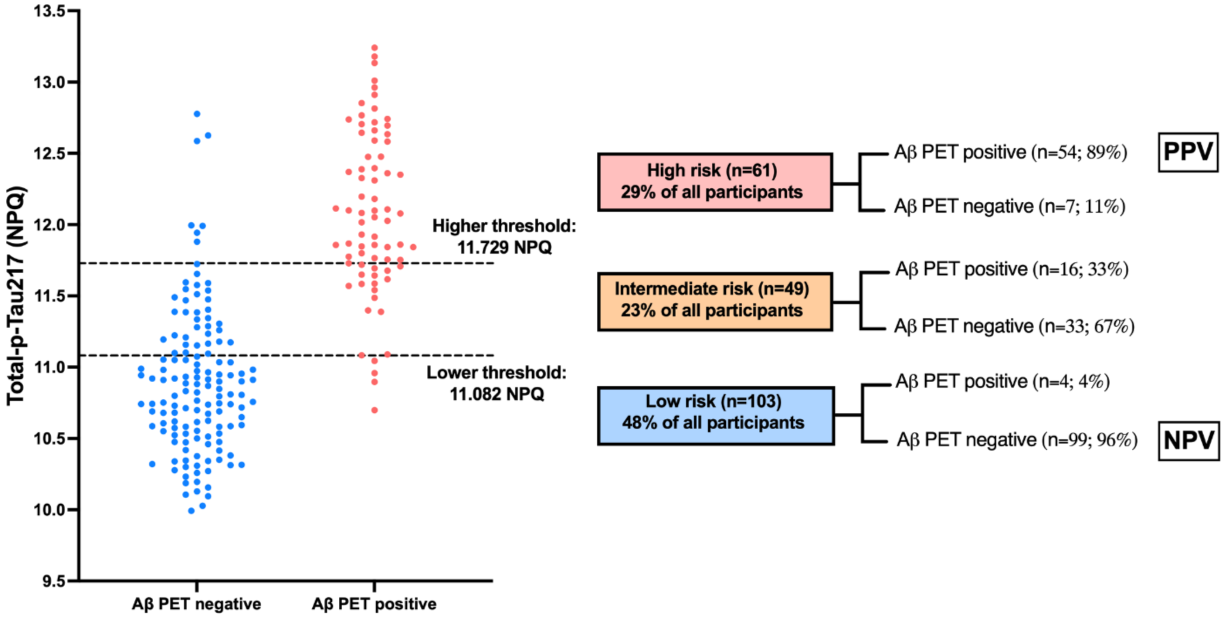


Three-range reference for Aβ-PET positivity. Distribution of plasma total-p-Tau217. The blue dots corresponded to individuals who are Aβ-PET negative and red dots to individuals who are Aβ-PET positive. In the three-range reference (A), the lower dashed line demonstrates where the 95% sensitivity low-risk threshold falls on the distribution, with the upper line corresponding to the 95% specificity High-risk threshold. On the right of each graph, the flowchart demonstrated the overall accuracy of the workflow.

Abbreviations: NPQ, NULISA protein quantification; NPV, negative predictive value; PET, positron emission tomography; PPV, positive predictive value

### Supplementary Figure S4. Prognostic performance of the plasma BD-p-Tau217-derived risk groups (based on binary reference)

#### (A) MMSE

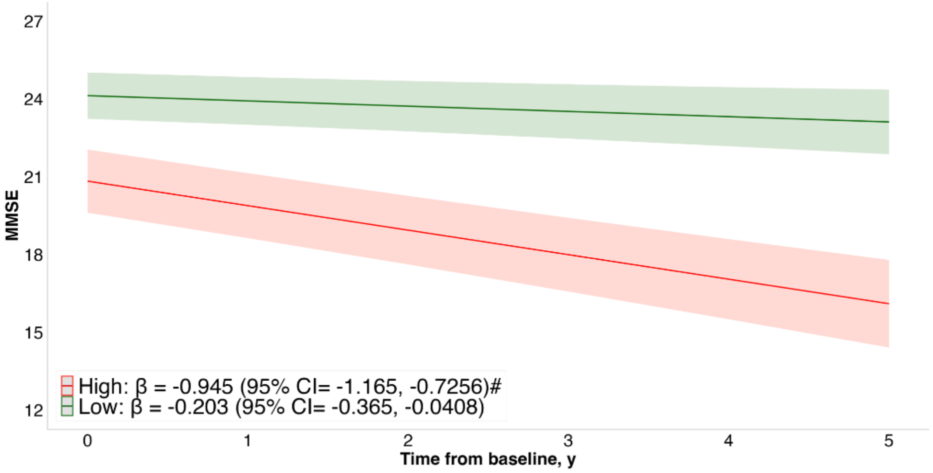


#### (B) CDR-SB

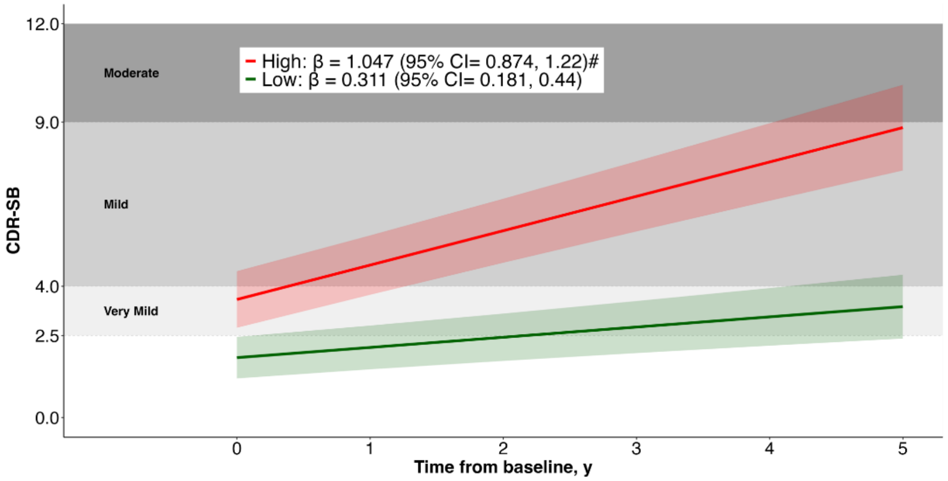


Associations between plasma BD-p-Tau217-derived risk groups with baseline and longitudinal cognitive performance. Trajectory plots indicate the mean longitudinal trajectories (solid line) of (A) MMSE and (B) CDR-SB, and associated 95% confidence intervals (shaded areas), estimated with linear-mixed effects models. Trajectories are stratified based on the plasma BD-p-Tau217-derived risk groups for PET Aβ positivity (Low-risk [green] vs High-risk [red]; derived from the binary reference), modeled with an interaction between risk groups and time. Models included random slopes and intercepts and were adjusted for age, sex, and years of education. For CDR-SB, shaded background regions indicate dementia severity categories based on CDR-SB thresholds from [1]: 2.5–4.0 (very mild), 4.5–9.0 (mild), 9.5–15.5 (moderate), and 16.0–18.0 (severe). #Indicates longitudinal trajectories that are significantly different from low-risk group.

Abbreviations: BD, brain-derived; MMSE, Mini-Mental State Examination; CDR-SB, Clinical Dementia Rating Sum of Boxes; PET, positron emission tomography

### Supplementary Figure S5. Kaplan-Meier survival curve for progression to dementia

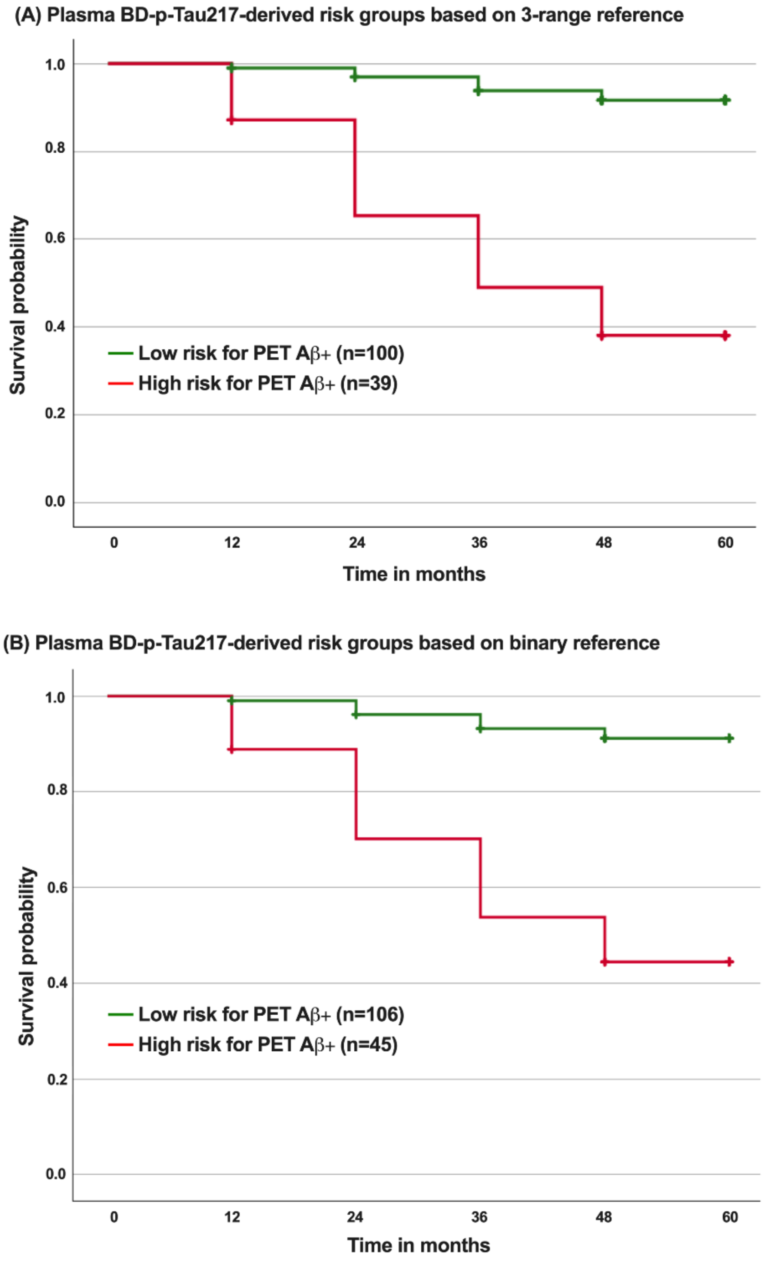


Kaplan-Meier survival curves reflecting progression to dementia among non-dementia participants (CN + CIND). Participants were stratified by risk of amyloid PET positivity (Low-risk versus High-risk), derived based on (A) the 3-range reference or (B) the binary reference of plasma BD-p-Tau217.

Abbreviations: BD, brain-derived; CIND, cognitive impairment no dementia; CN, cognitively normal; PET, positron emission tomography

**Reference**

1. O'Bryant, S.E., et al., *Validation of the new interpretive guidelines for the clinical dementia rating scale sum of boxes score in the national Alzheimer's coordinating center database.* Arch Neurol, 2010. **67**(6): p. 746-9.
